# Prevalence of Dengue and Chikungunya in the Horn of Africa: A Systematic Review and Meta-analysis

**DOI:** 10.64898/2026.08.03.26359549

**Authors:** Aamir M. Osman, Mercy Mwenya, Francis Ochieng, Hellen Akengo, Joel Lutomiah, Konongoi Limbaso, Ahmed Abdulkadir Hassan-Kadle, Fredrick Ouma Odhiambo, Marian Muse Osman, Bernard Bett

## Abstract

**Background:** Dengue virus (DENV) and Chikungunya virus (CHIKV) are important mosquito-borne arboviruses with expanding global distributions. Although recurrent outbreaks have occurred in the Horn of Africa since the early 2000s, their epidemiology and burden remain poorly characterized. This systematic review and meta-analysis estimated the prevalence of DENV and CHIKV, their distribution by country, diagnostic methods, circulating DENV serotypes, and mosquito vectors.

**Methods:** Following PRISMA guidelines, PubMed, Web of Science, Scopus, SpringerLink, Nature Portfolio, and Google Scholar were searched for studies published up to October 2025. Eligible studies reported DENV and/or CHIKV infections in humans or mosquitoes in Kenya, Ethiopia, or Somalia. Random-effects meta-analyses were performed, with subgroup analyses by country, population, setting, and diagnostic method.

**Results:** Of 3,351 records, 65 studies met the inclusion criteria, most from Kenya (63.1%), followed by Ethiopia (30.8%) and Somalia (6.1%). Pooled DENV prevalence was 20% for IgG, 10% for IgM, 6% for NS1, and 4% by PCR. Corresponding CHIKV prevalence was 5% for IgG, 8% for IgM, and 4% by PCR. Significant differences between countries were observed for DENV NS1 (p<0.0001) and PCR-confirmed infections (p=0.01). DENV prevalence was highest in the general population (16%) compared with febrile patients (6%) and mosquito vectors (1%) (p=0.033), whereas CHIKV prevalence was highest in rural settings (15%) (p=0.0002). All four DENV serotypes circulated in Kenya and Ethiopia, while DENV-2 and DENV-3 were reported in Somalia. *Aedes aegypti* was the predominant vector, reported in 92.3% of vector studies.

**Conclusions:** DENV and CHIKV are important and likely underrecognized arboviral threats to global public health and in the Horn of Africa, with evidence of ongoing transmission and widespread exposure. Strengthened surveillance, diagnostic capacity, vector control, and regional collaboration are needed to mitigate their growing public health impact.

**Author summary:** Dengue and chikungunya are viral diseases transmitted by mosquitoes and are responsible for an increasing number of outbreaks worldwide. In the Horn of Africa, these infections are often missed because their symptomatology mimics malaria and other common febrile illnesses and laboratory testing is limited. We reviewed all published studies available from Kenya, Ethiopia and Somalia to better understand the epidemiology, geographic distribution, mosquito vectors and evidence for virus circulation in the region. We found widespread exposure to both viruses, evidence of ongoing transmission, and multiple circulating dengue virus types in Kenya and Ethiopia. We also found that the principal mosquito responsible for the transmission of these viruses world-wide, *Aedes aegypti*, was the most reported vector throughout the region. However, our review identified significant gaps in surveillance, particularly in Somalia and parts of Ethiopia, limited laboratory diagnostic capacity and few studies on mosquito ecology. These observations suggest that the actual burden of dengue and chikungunya is underestimated. We believe that improved outbreak detection, preparedness and disease control in the Horn of Africa will result from better surveillance, improved diagnostic capacity, wider mosquito surveillance and increased regional collaboration via a One Health approach.

## Introduction

Mosquito-borne arboviruses represent an increasing global public health threat, particularly in tropical and subtropical regions where environmental conditions favor their vectors’ proliferation [1,2]. Among these, dengue virus (DENV), a member of the genus *Flavivirus*, and chikungunya virus (CHIKV), belonging to the genus *Alphavirus*, are two of the most widespread and rapidly expanding arboviruses worldwide [3]. Both viruses are primarily transmitted by *Aedes* mosquitoes, especially *Aedes aegypti* and *Aedes albopictus*, and are responsible for substantial morbidity, recurrent outbreaks, and growing pressure on healthcare systems [2,3].

The global burden of DENV and CHIKV has increased substantially over the recent decades. Dengue is currently the most widespread mosquito-borne viral disease, causing an estimated 400 million infections and approximately 36,000 deaths annually [4]. Similarly, CHIKV has re-emerged as a major public health concern, with recurrent outbreaks reported across tropical and sub-tropical regions, resulting in considerable health and economic burdens in affected populations [5]. The expansion of both viruses has been driven by rapid urbanization, increased human mobility, population growth, environmental change, and inadequate public health infrastructure [6]. In addition, climate change has enhanced the transmission potential of the primary vectors, *Aedes aegypti* and *Aedes albopictus*. Between 1951–1960 and 2015–2024, climate suitability for dengue transmission increased by 11.6% for *Ae. aegypti* and 48.5% for *Ae. albopictus*, contributing to the unprecedented rise in dengue cases reported globally in recent years [7].

Although arboviral diseases have historically received less attention in Africa than other vector-borne diseases such as malaria, growing evidence indicates widespread circulation of DENV and CHIKV across the continent. East Africa, in particular, provides favorable ecological conditions for transmission due to its tropical and subtropical climate, rapid urban expansion, and widespread distribution of competent mosquito vectors [8,9]. Recurrent outbreaks and serological surveys have demonstrated sustained circulation of both viruses in several countries within the region.

The Horn of Africa, including Kenya, Ethiopia, and Somalia, has experienced multiple outbreaks of dengue and chikungunya over the past two decades [10]. Kenya has reported recurrent dengue and chikungunya outbreaks, particularly in coastal and northeastern regions [11,12], while Ethiopia has experienced repeated dengue outbreaks since 2013 and documented chikungunya transmission since 2016 [13]. In Somalia, dengue cases have been reported since the early 1990s, with increasing evidence of continued transmission and periodic outbreaks [14]. Together, these findings suggest that both viruses are becoming increasingly established within the region.

The co-circulation of DENV and CHIKV presents important public health challenges. Both infections share similar clinical manifestations, including acute fever, headache, myalgia, and rash, which frequently results in misdiagnosis, particularly in malaria-endemic settings [15]. While chikungunya is often characterized by debilitating arthralgia, severe dengue can lead to hemorrhagic complications and increased mortality [11]. Limited diagnostic capacity, under-resourced healthcare systems, and weak surveillance programs further contribute to the under recognition and underreporting of these infections [16,17]. Furthermore, the circulation of multiple DENV serotypes is of particular concern because secondary infection with a heterologous serotype may increase the risk of severe disease. Understanding the distribution of circulating serotypes and mosquito vectors is therefore essential for strengthening outbreak preparedness and control strategies.

Despite increasing reports of DENV and CHIKV transmission in the Horn of Africa, the available evidence remains fragmented and is largely derived from outbreak investigations, localized serosurveys, and hospital-based studies. Consequently, the regional burden, transmission dynamics, serotype distribution, and vector ecology of these arboviruses remain poorly characterized. A comprehensive synthesis of the available evidence is therefore needed to better understand the epidemiology of dengue and chikungunya and to identify gaps or research questions that future studies should address.

Therefore, this systematic review and meta-analysis aimed to estimate the pooled serological and molecular prevalence of DENV and CHIKV in the Horn of Africa, assess variations in prevalence by country, population group, study setting, and diagnostic method, and describe the distribution of DENV serotypes and reported mosquito vectors in the region. It also synthesizes the findings generated to identify new leads for future work.

## 2. Materials and Methods

### 2.1 Methods Reporting and protocol registration

This systematic review and Meta-analysis was conducted in accordance with the Preferred Reporting Items for Systematic reviews and Meta-Analyses (PRISMA) guidelines [18], to ensure transparency, reliability, and traceability of the study through consistent reporting. The protocol for this review was registered with the International Platform of Registered Systematic Review and Meta-analysis Protocols INPLASY (Unique ID number: INPLASY202640055) and is available in full on inplasy.com (DOI: https://doi.org/10.37766/inplasy2026.4.0055).

### 2.2 Data sources and search strategies

To find reliable and relevant studies on the epidemiology of dengue and chikungunya within the Horn of Africa, several electronic databases for scientific papers were extensively searched. The databases include PubMed, Web of Science, Scopus, SpringerLink, Nature Portfolio, and Google Scholar. The search aimed to identify peer-reviewed published scientific papers from studies conducted in Kenya, Ethiopia, and Somalia that reported on dengue and chikungunya viruses. The search was conducted by October 8, 2025.

Search terms were developed based on the review objectives and combined using Boolean operators (“AND” and “OR”) and truncation () where appropriate. The primary search string included: (“dengue” OR “dengue virus” OR “DENV” OR “chikungunya” OR “chikungunya virus” OR “CHIKV”) AND (“Aedes” OR “vector*” OR “mosquito*”) AND ((“Kenya” OR “Somalia” OR “Ethiopia” OR “East Africa” OR “Horn of Africa” OR “Western Kenya”) OR (“Mombasa” OR “Nairobi” OR “Kisumu” OR “Eldoret” OR “Nyanza” OR “Mandera” OR “Garisa” OR “Coast*” OR “Kilifi” OR “Kwale” OR “Isiolo” OR “Wajir”) OR (“Addis Ababa” OR “Oromi*” OR “Amhara” OR “Tigray” OR “Dire Dawa” OR “Somali Region”) OR (“Mogadishu” OR “Hargeisa” OR “Kismayo” OR “Puntland” OR “Somaliland” OR “Jubaland”).

### 2.3 Study selection

All records identified through the database searches were imported into Rayyan, an AI-assisted systematic review management platform, to facilitate study organization and screening. Duplicate records were identified and removed prior to screening. Study selection was conducted according to the predefined inclusion and exclusion criteria in a two-stage process. First, two independent reviewers (AMO and MM) screened titles and abstracts to identify potentially eligible studies. Following this initial screening, 77 articles were selected for full-text assessment. Subsequently, full-text screening was performed, resulting in the inclusion of 65 studies that met the eligibility criteria. Additional reviewer (BB) provided oversight throughout the screening process and assisted in resolving discrepancies. Any disagreements at either stage of screening were resolved through discussion and consensus among the reviewers.

### 2.4. Inclusion criteria

Studies were eligible for inclusion if they reported on DENV and/or CHIKV infection, outbreaks, surveillance, detection, molecular characterization, or serotype distribution in human and/or mosquito vector populations. Only studies conducted in Kenya, Ethiopia, or Somalia were considered. Relevant outcomes included prevalence, incidence, molecular detection, virus isolation, serological evidence of infection, outbreak investigations, and genetic characterization of DENV or CHIKV. Observational studies, including cross-sectional, case-control, cohort, and surveillance studies, as well as laboratory-based and field-based investigations involving human or vector samples, were eligible for inclusion. Only articles published in English and subjected to peer review were included.

### 2.5. Exclusion criteria

Studies were excluded if they were conducted outside Kenya, Ethiopia, or Somalia; focused on arboviruses other than DENV or CHIKV; or investigated vector competence unrelated to dengue or chikungunya transmission. Reviews, editorials, commentaries, conference abstracts, and other publications lacking original data were also excluded. Additionally, studies that did not report relevant outcomes, such as prevalence, detection, outbreak data, or molecular characterization, were excluded from the review.

### 2.6. Data extraction

A standardized data extraction form was developed using Microsoft Excel^®^ 365 (Microsoft Corporation, Redmond, WA, USA) to ensure consistent collection of relevant information from eligible studies. Two reviewers (AMO and MM) independently extracted data from each included full-text article, and any discrepancies were resolved through discussion and consensus. Extracted information included publication details (year and month), study location, study objectives, target population, sampling design and sample size, diagnostic methods used, and reported outcomes related to DENV and CHIKV, including prevalence, serotypes, genotypes, outbreaks, and mortality. Information on vector species and study limitations was also collected where available.

### 2.7. Quality assessment

The critical appraisal for the selected studies was done to assess the quality of the study and the reliability of the data generated. It was carried out by two independent reviewers (AMO and MM). For this reason, the Joanna Briggs Institute (JBI) critical appraisal checklist for cross-sectional studies was used [19]. The checklist consists of eight criteria that analyse the quality of the paper. The assessment analysed the clarity of the sample and setting description, risk factors definition, identified confounders, measurement of disease outcome, and the appropriateness of the statistical methods used, if any. The answers permitted were “Yes”, “No”, “Unclear” and “Not applicable”, which each had an assigned value. Only studies that met the minimum quality mark of 50% or higher were included, and all the studies met the criteria. The scores for this were recorded in a Microsoft Excel sheet.

### 2.8. Data analysis

Data were extracted into Microsoft Excel and analyzed using R software. Descriptive analyses were conducted to summarize study characteristics, including country, population, study setting, diagnostic methods, and reported vector species. Meta-analyses of prevalence were performed using the meta package in R. Pooled prevalence estimates for DENV and CHIKV were calculated using a random-effects model to account for between-study variability. Proportions were transformed using the logit transformation (PLOGIT) to stabilize variances. The inverse-variance method was used for weighting individual studies.

Heterogeneity across studies was assessed using the I² statistic, with values above 75% considered indicative of substantial heterogeneity. Subgroup analyses were conducted to explore potential sources of heterogeneity based on diagnostic method (e.g., serology vs molecular), study setting (urban, peri-urban, rural), population type (general population, febrile patients, and vectors), and country. Differences between subgroups were evaluated using the chi-square test for subgroup differences. All statistical analyses were conducted at a significance level of *P <* 0.05.

## 3. Results

### 3.1 Literature selection

The study selection process is presented in Figure 1. A total of 3,351 records were identified through database searches. After removing 1,149 duplicates and one retracted article, 2,202 records remained for title and abstract screening, of which 2,127 were excluded. Subsequently, 73 full-text articles were assessed for eligibility (two could not be retrieved), and eight studies were excluded due to wrong outcome (n = 4), wrong publication type (n = 3), or inappropriate study design (n = 1). Ultimately, 65 studies were included in the systematic review.

**Figure 1.**
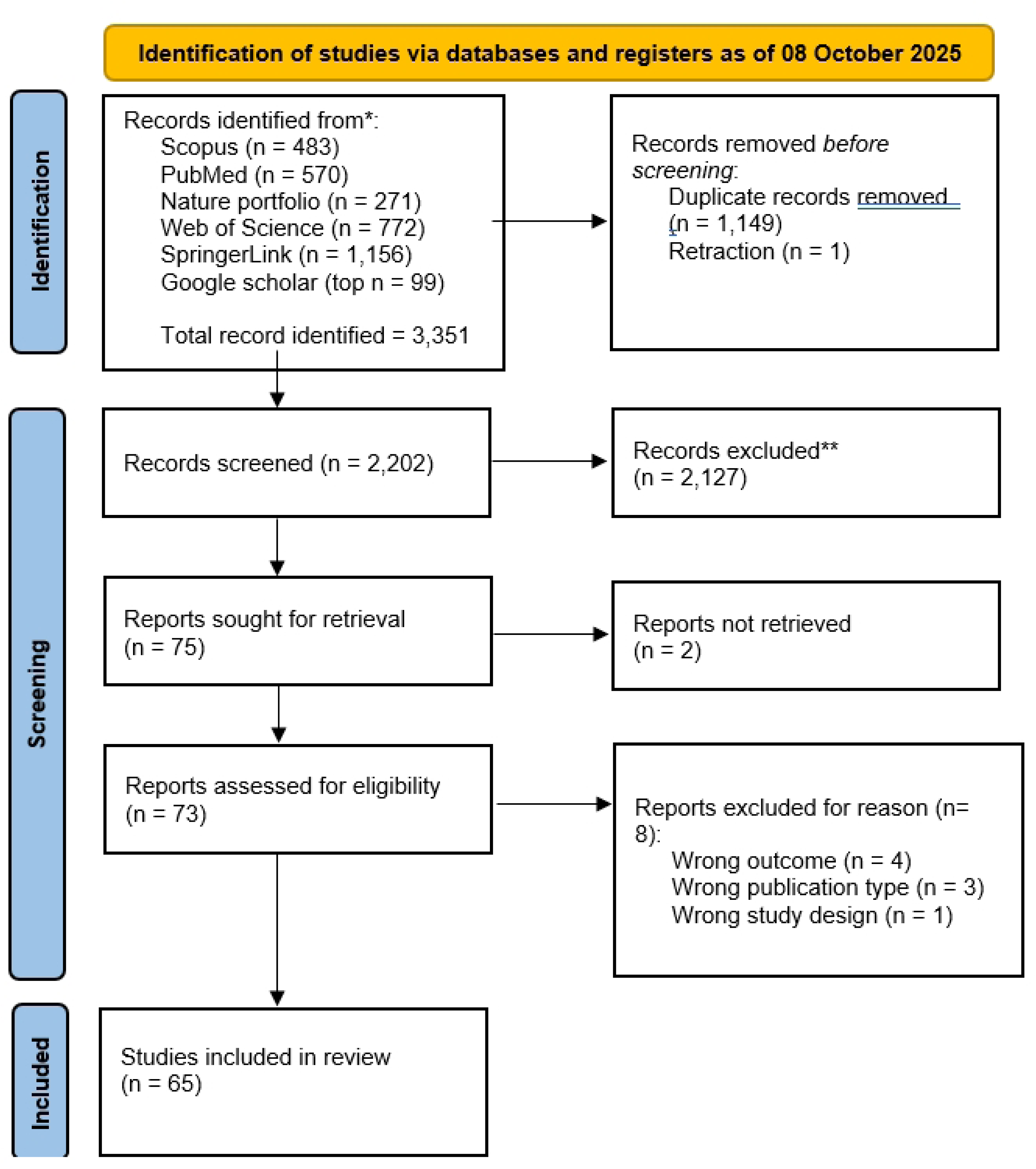
PRISMA 2020 flow diagram illustrating the study selection process for the systematic review.

### 3.2 Assessment of publication quality

The methodological quality of the included studies, assessed using the Joanna Briggs Institute critical appraisal (JBI) tool, ranged from 75% to 100% across applicable domains, with a mean score of 95.3% (Supplementary table S1). Most studies clearly defined eligibility criteria described study populations and settings and used valid outcome measurement methods. All studies met acceptable quality thresholds and were classified as low risk of bias; therefore, none were excluded based on quality assessment.

### 3.3 Characteristics of the included studies

A total of 65 studies published between 1998 and 2025 were included. Most studies were conducted in Kenya 41/65 (63.1%), followed by Ethiopia 20/65 (30.8%) and Somalia 4/65 (6.1%) (Figure 2). Based on the extracted disease indicators, DENV was the most frequently investigated arbovirus 31/65 (47.6%) studies, followed by CHIKV 14/65 (21.5% studies). Other arboviruses that were studied together with these two were yellow fever and Zika virus, which were reported in 30 studies. Nineteen studies (19/65, 29.2% studies) investigated both DENV and CHIKV (Figure 3).

**Figure 2.**
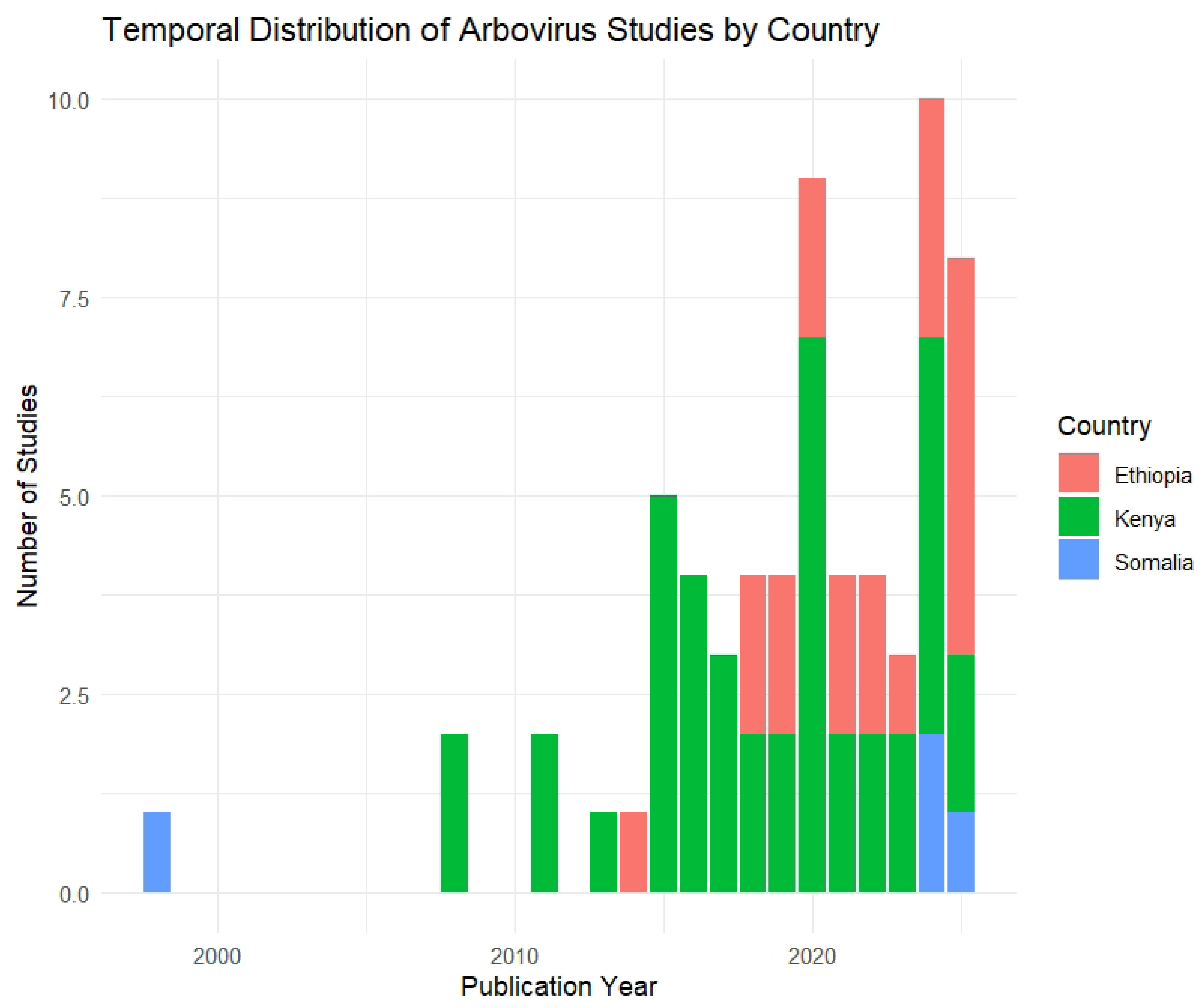
Geographic distribution of the 65 included studies by country conducted between 1998 and 2025.

**Figure 3.**
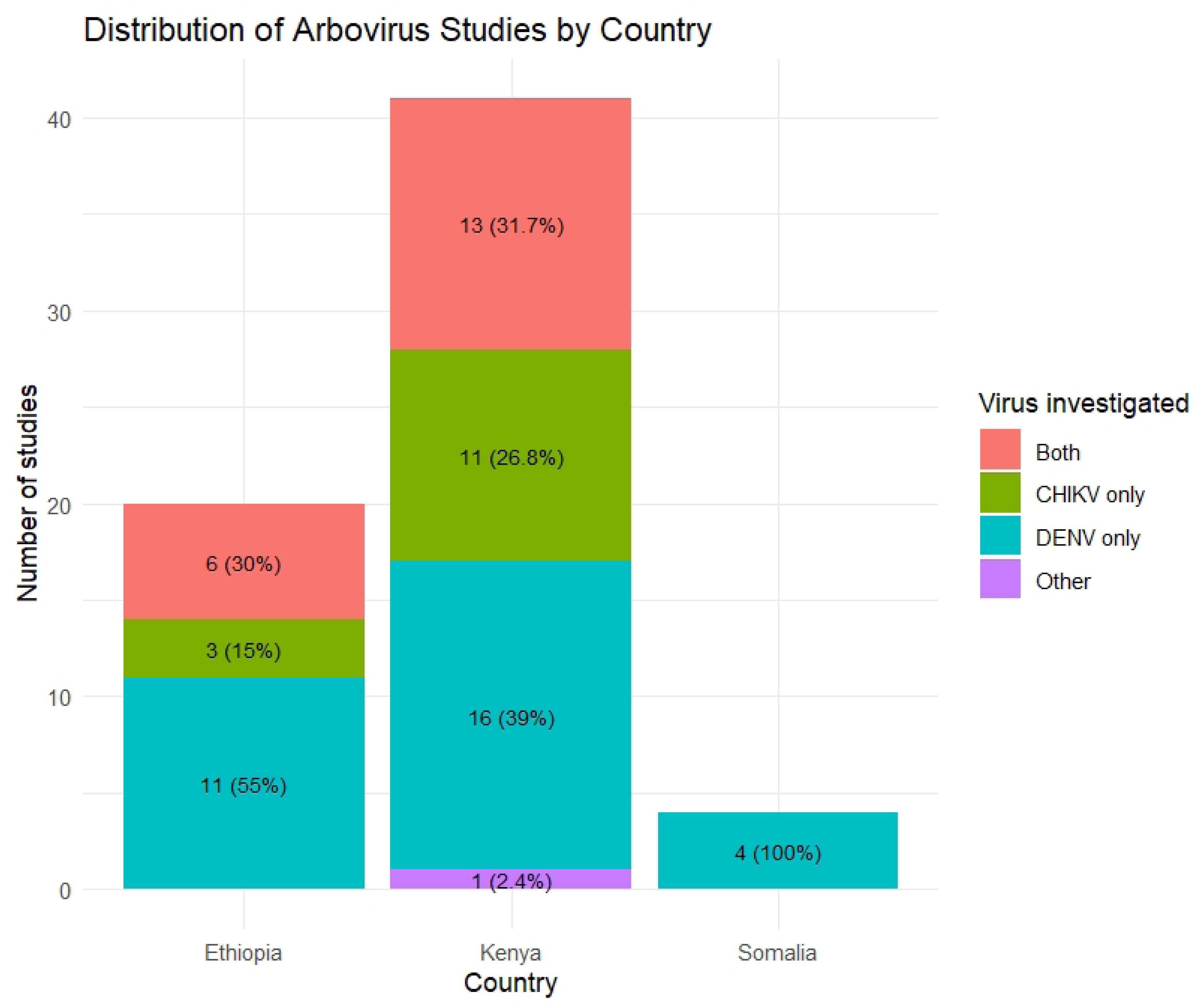
Distribution of arboviruses investigated in the included studies.

Regarding diagnostic approaches, molecular and serological methods were most used, particularly serology-based assays (40/65, 61.5%), and molecular (35/65, 53.8%). The most frequent target population was febrile patients (30/65, 46.1%), followed by apparently healthy population, referred here as general population samples (13/65, 20%) and mosquito vectors (9/65, 13.8%). Studies were conducted across rural (24/65, 36.9%), urban (21/65, 32.3%), and peri-urban (20/65, 30.8%) settings. Most studies used a cross-sectional sampling design (34/65, 52.3%), with fewer retrospective (9/65, 13.8%) and longitudinal designs (5/65, 7.7%) (Figure 4).

**Figure 4.**
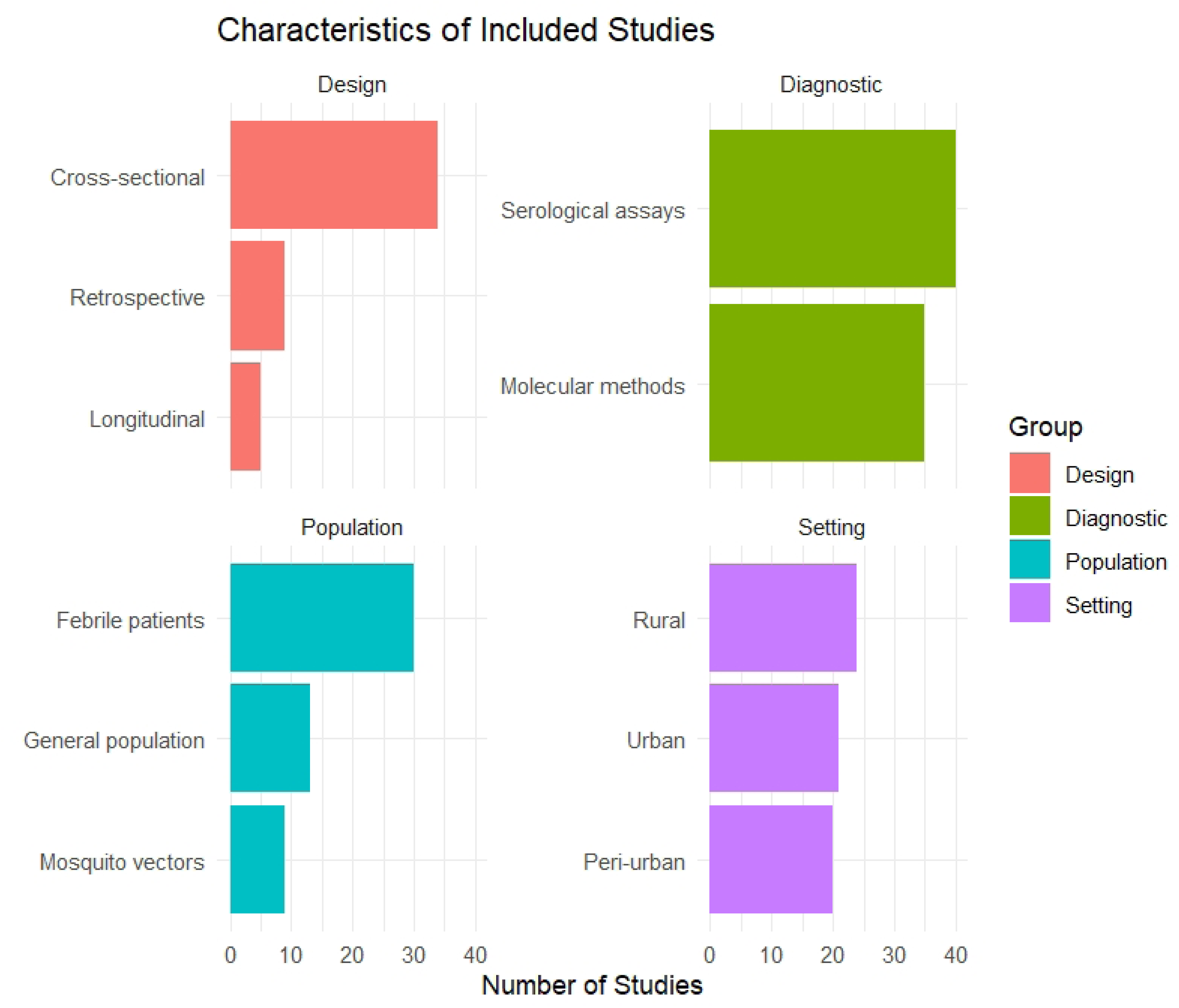
Methodological characteristics of the included studies.

### 3.4 Dengue virus serological and molecular prevalence

A total of 24,713 participants across multiple studies were included in the meta-analyses of DENV infection markers. The pooled DENV IgG seroprevalence was 20% (95% CI 12–33%). Considerable heterogeneity was observed (I² = 99.5%, *P <* 0.0001). Subgroup analysis showed similar pooled estimates in Ethiopia (21%; 95% CI 8–46%) and Kenya (18%; 95% CI 9–33%), with no significant difference between countries (*P* = 0.79) (Figure 5). The pooled DENV IgM prevalence was 10% (95% CI 6–17%). Heterogeneity remained high (I² = 96.9%, *P <* 0.0001).

**Figure 5.**
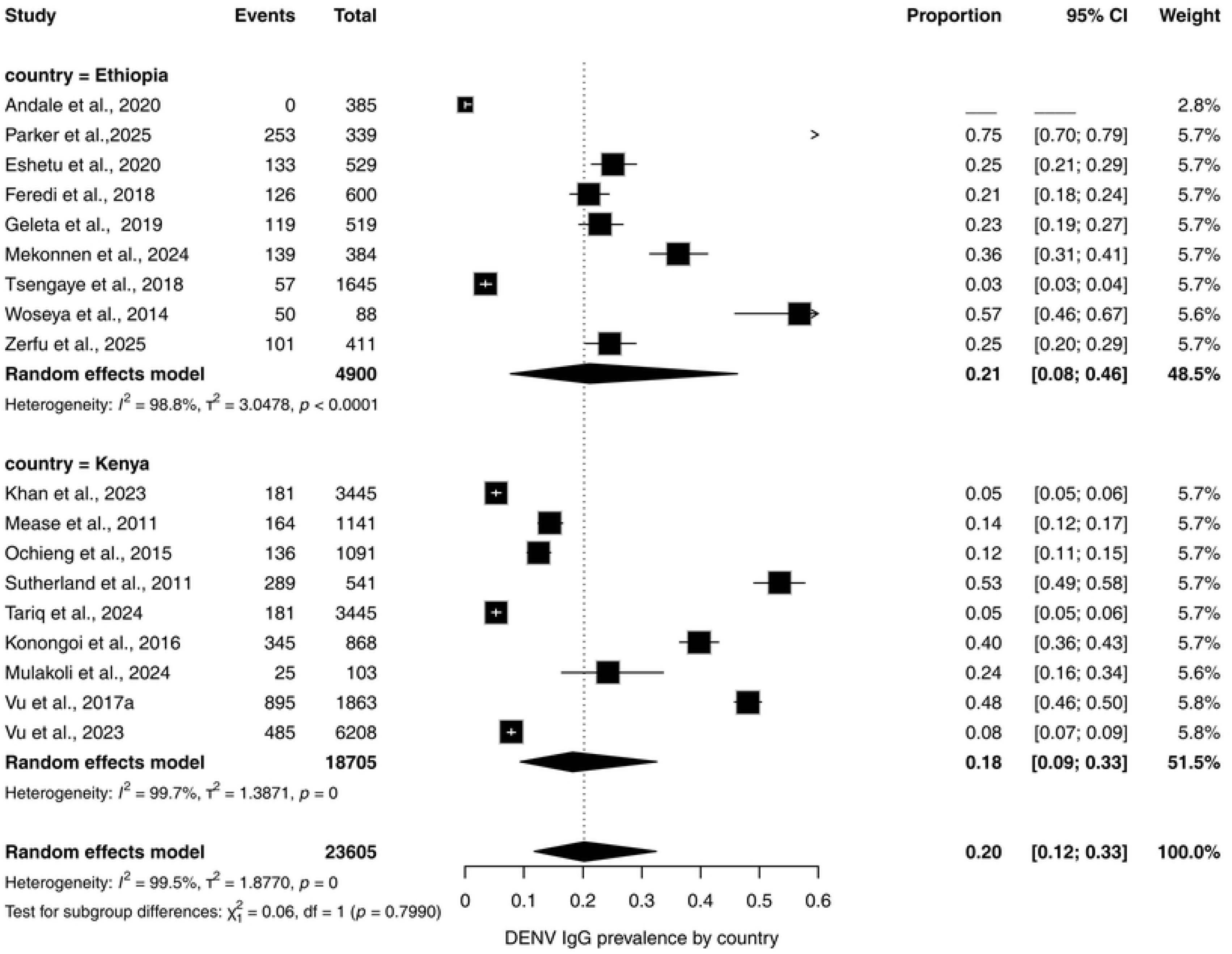
Forest plot showing the pooled prevalence of dengue virus IgG antibodies with subgroup analysis by country.

Country-specific estimates were 17% (95% CI 10–29%) in Ethiopia, 12% (95% CI 8–19%) in Kenya, and 4% (95% CI 1–16%) in Somalia, although between-country differences were not statistically significant (*P* = 0.14) (Figure 6).

**Figure 6.**
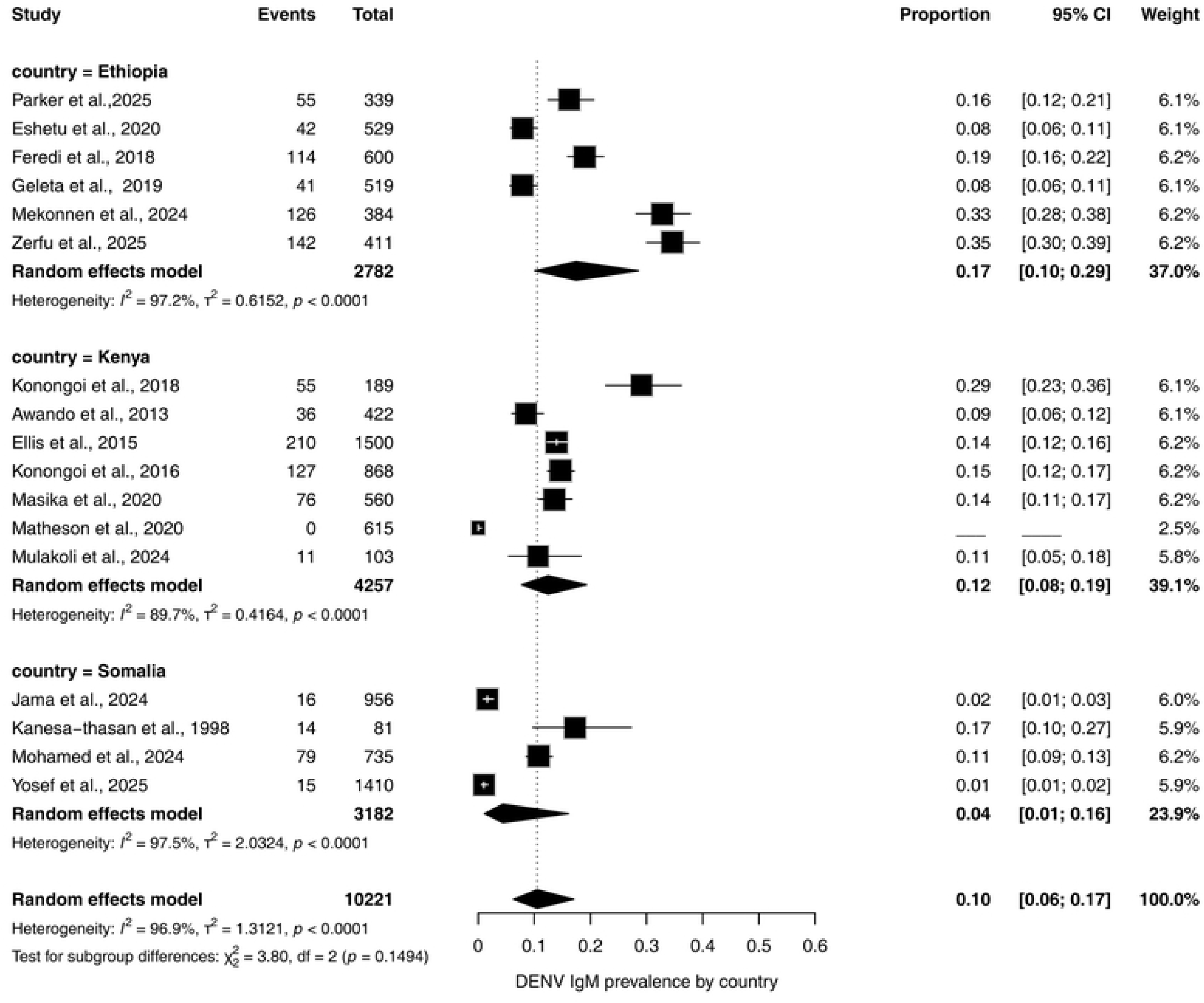
Forest plot showing the pooled prevalence of dengue virus IgM antibodies with subgroup analysis by country.

Regarding DENV NS1 antigen, four studies assessed provided a pooled prevalence of 6% (95% CI 3–12%). Substantial overall heterogeneity was observed (I² = 84.9%). Somalia demonstrated the highest pooled NS1 prevalence at 11% (95% CI 9–12%), compared with 5% (95% CI 1–18%) in Kenya and 1% in Ethiopia. Between-country differences were statistically significant (*P <* 0.0001) (Figure 7). PCR-confirmed DENV prevalence was 4% (95% CI 2–10%). Heterogeneity was considerable (I² = 96.2%, *P <* 0.0001). Pooled estimates were 7% (95% CI 2–20%) in Ethiopia, 3% (95% CI 1–10%) in Kenya, and 17% in Somalia (single study). Subgroup differences were statistically significant (*P* = 0.01) (Figure 8).

**Figure 7.**
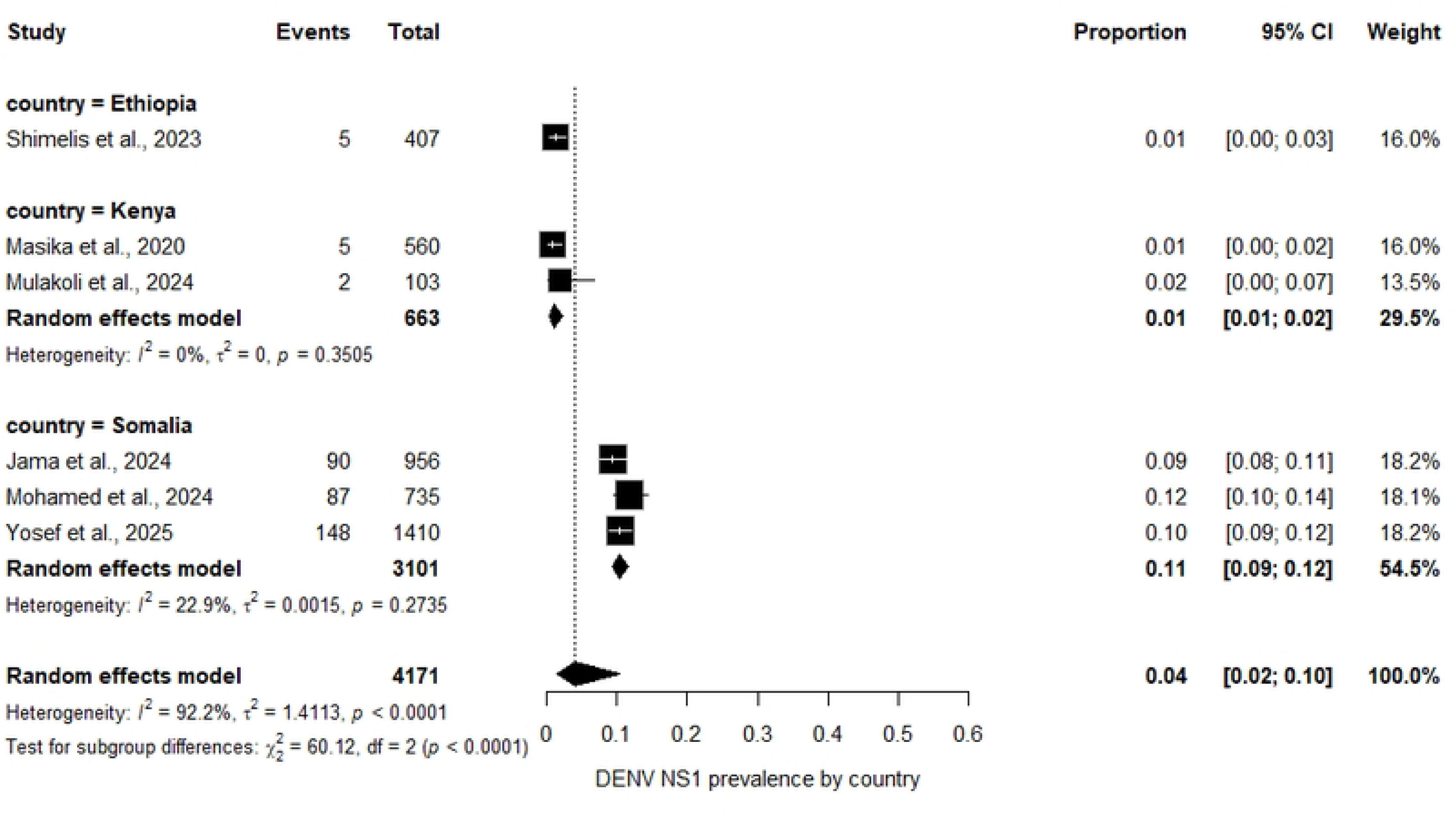
Forest plot showing the pooled prevalence of dengue virus NS1 antigen with subgroup analysis by country.

**Figure 8.**
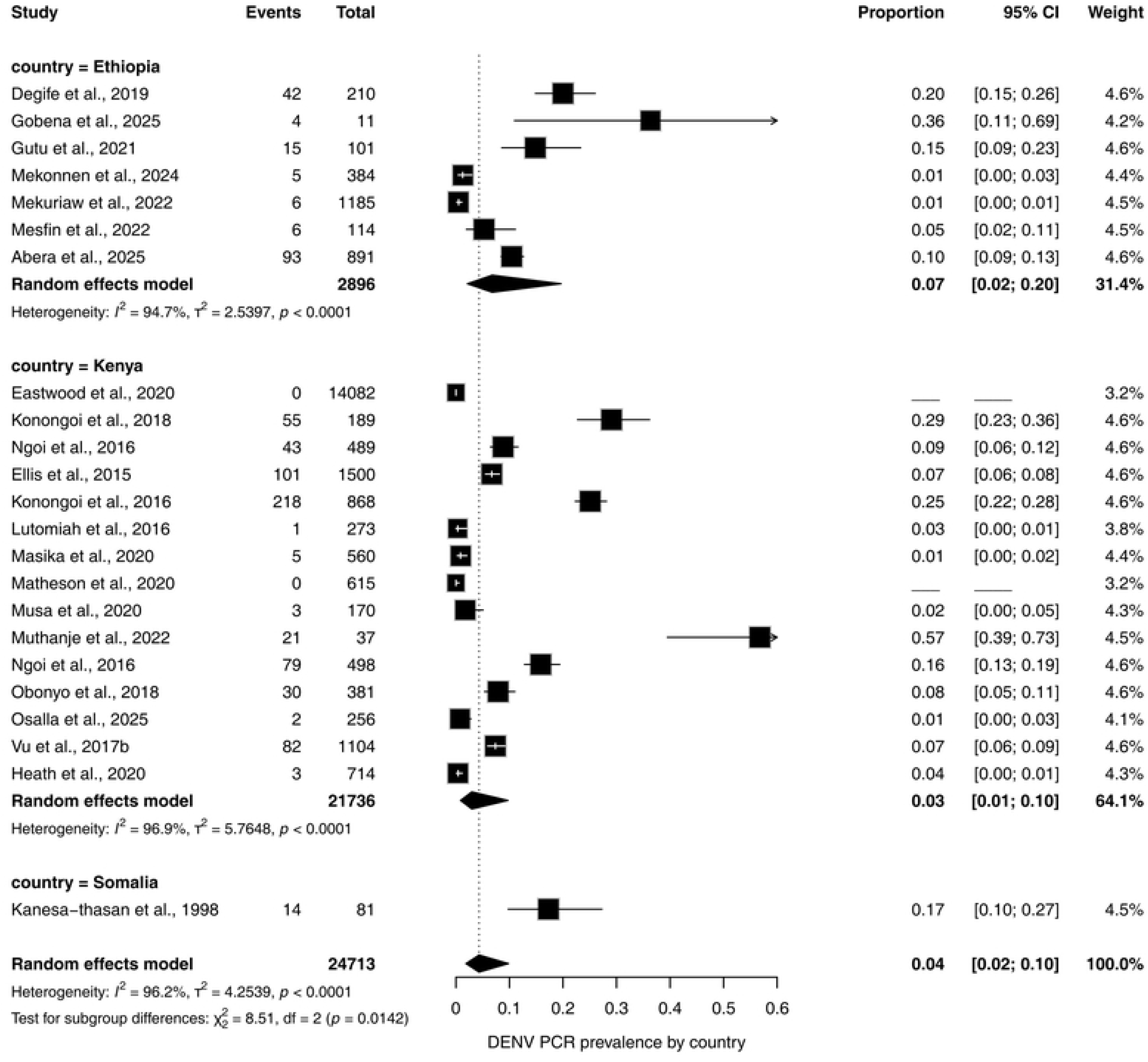
Forest plot showing the pooled prevalence of PCR-confirmed dengue virus infection with subgroup analysis by country.

### 3.5 DENV prevalence by subgroup analyses

Subgroup analyses revealed varying patterns of DENV prevalence across diagnostic methods, settings, and population groups. Studies employing combined serological and molecular approaches reported a pooled prevalence of 6%, compared to 4% in studies using molecular methods alone; however, this difference was not statistically significant (χ² = 0.34, *P* = 0.5617) (Figure 9). Similarly, prevalence varied by study setting, with higher estimates observed in urban (7%) and peri-urban areas (6%) compared to rural settings (1%), though these differences were also not statistically significant (χ² = 2.55, *P* = 0.2797) (Figure 10). In contrast, population-based subgroup analysis demonstrated a significant difference (χ² = 6.82, *P* = 0.0330), with the highest prevalence observed in the general population (16%), followed by febrile patients (6%), and the lowest in mosquito vectors (1%). Across all subgroup analyses, substantial heterogeneity persisted (I² > 90%) (Figure 11).

**Figure 9.**
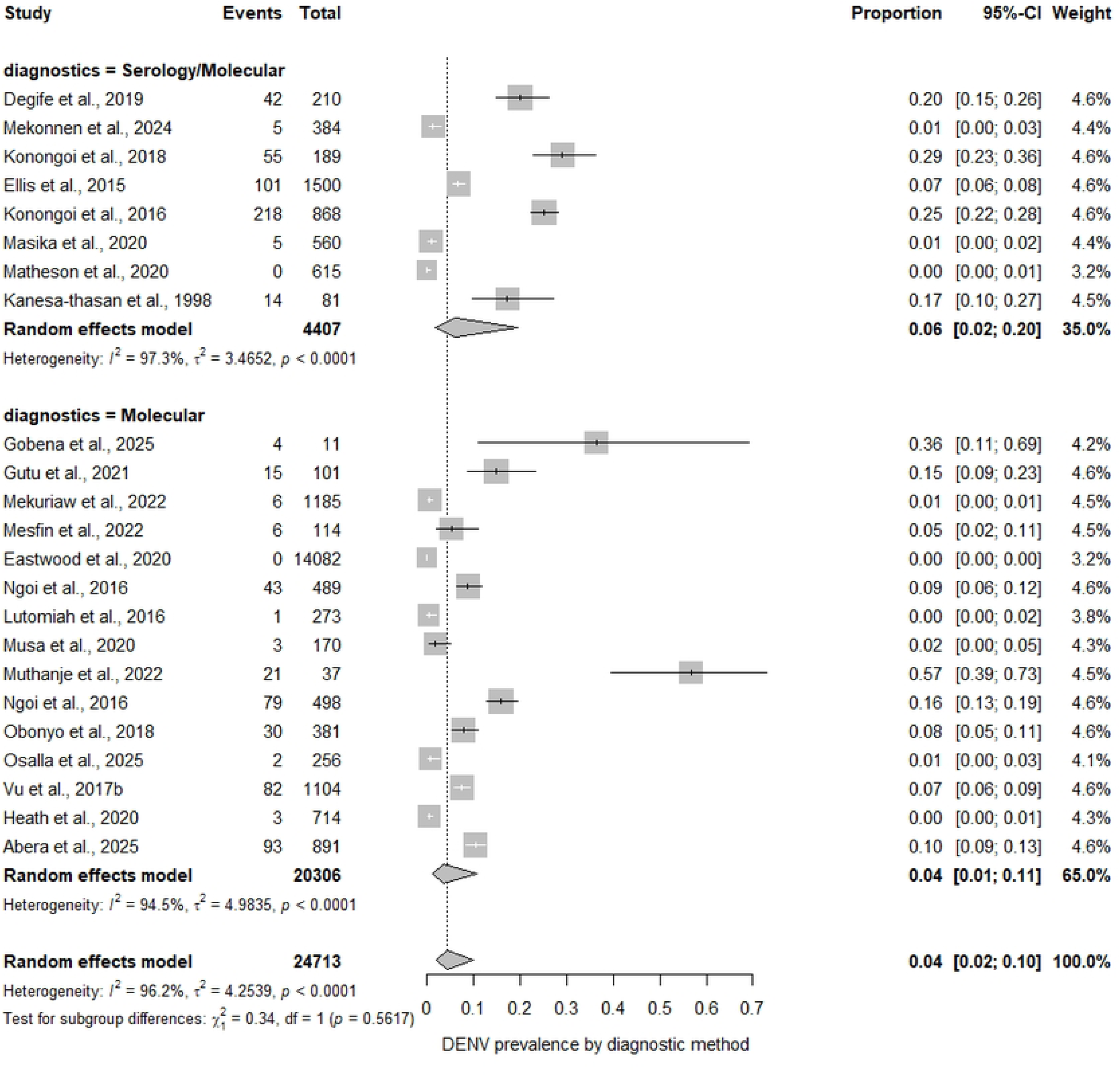
Subgroup analysis of PCR-confirmed dengue virus prevalence according to diagnostic approach.

**Figure 10.**
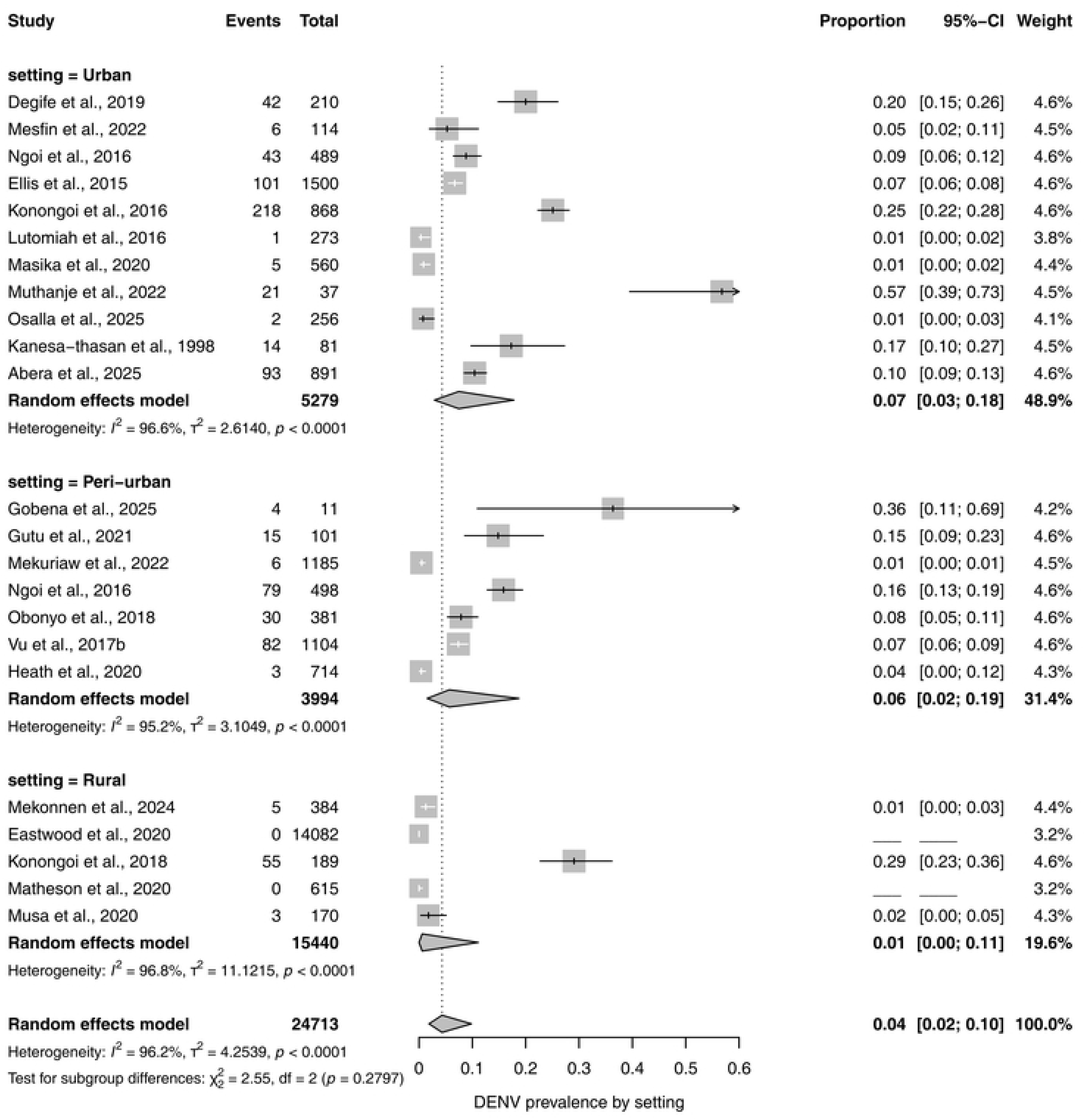
Subgroup analysis of PCR-confirmed dengue virus prevalence according to study setting.

**Figure 11.**
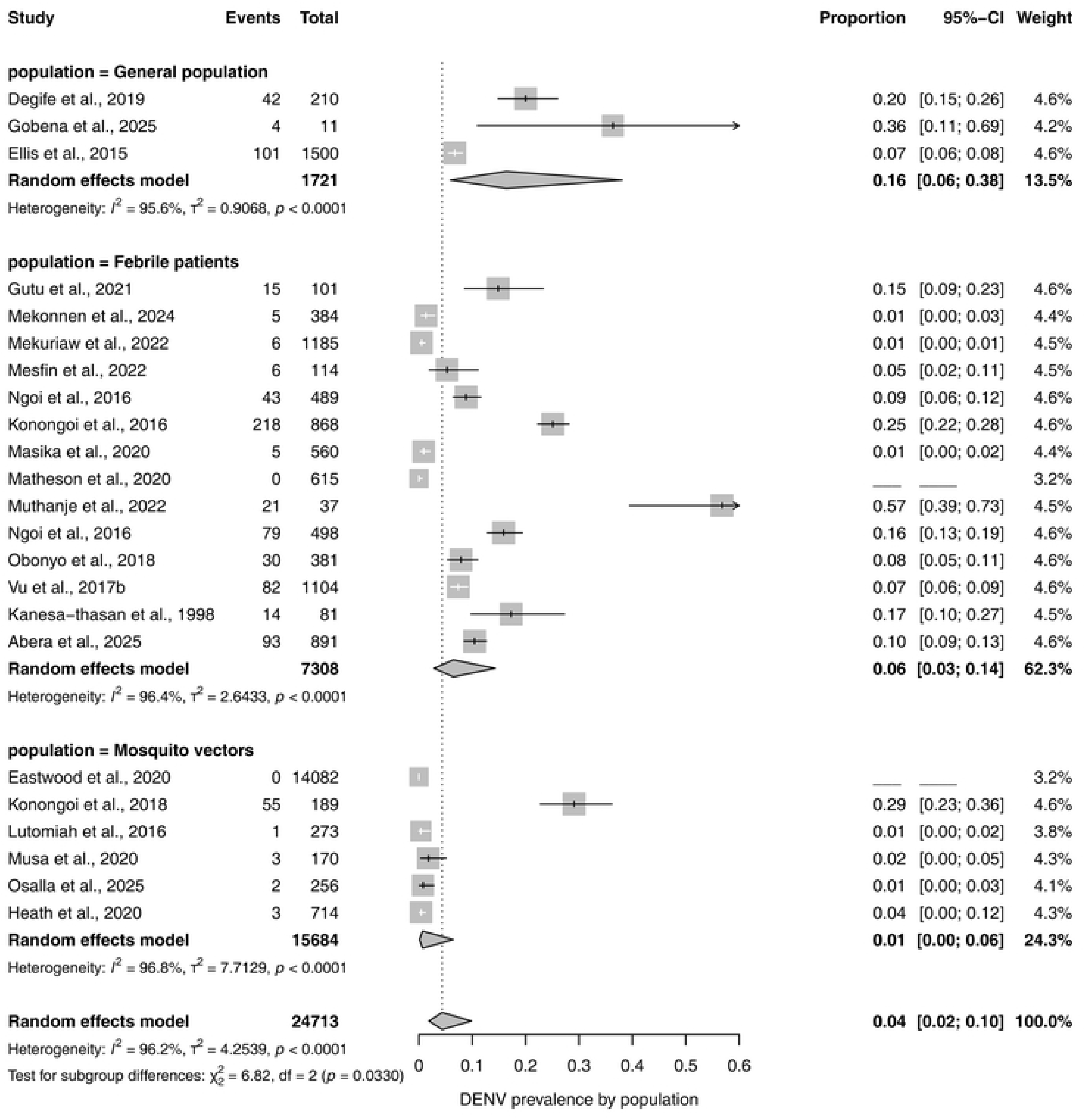
Subgroup analysis of PCR-confirmed dengue virus prevalence according to study population.

### 3.6 Distribution of dengue virus serotypes

The geographical distribution of DENV serotypes reported in the included studies is illustrated in (Figure 12). Across the included studies, Kenya showed the highest diversity and number of reports, with all four dengue serotypes identified, including DENV-2 in 14 studies, DENV-3 in 6 studies, DENV-1 in 4 studies, and DENV-4 in 2 studies. In Ethiopia, all four serotypes were also reported but with fewer studies, including DENV-3 in 5 studies, DENV-2 in 3 studies, DENV-1 in 2 studies, and DENV-4 in 1 study. In contrast, Somalia had more limited evidence, with only two serotypes detected, namely DENV-2 and DENV-3, each reported in one study.

**Figure 12.**
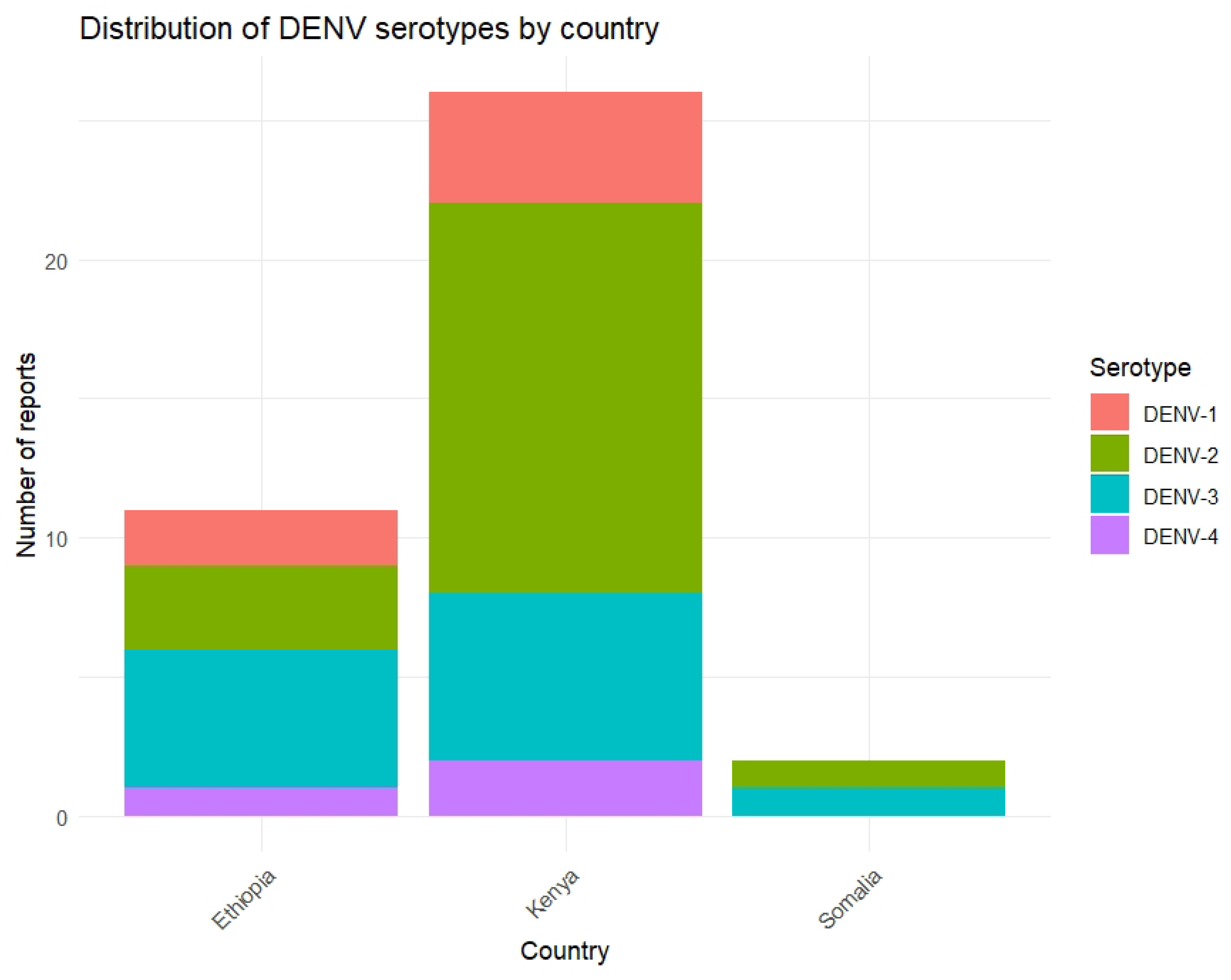
Geographic distribution of dengue virus serotypes reported across Kenya, Ethiopia, and Somalia.

### 3.7 Chikungunya virus serological and molecular prevalence

The pooled overall CHIKV IgG seroprevalence was 5% (95% CI 1–16%; I² = 99.1%). Country-specific estimates were 1% in Ethiopia and 11% in Kenya (Figure 13). The pooled overall CHIKV IgM prevalence was 8% (95% CI 2–30%; I² = 95.8%). Subgroup analysis showed higher estimates in Ethiopia (29%) compared to Kenya (4%), though heterogeneity remained considerable (I² = 98.4%) (Figure 14). For PCR-confirmed CHIKV, the overall pooled prevalence was 4% (95% CI 1–13%; I² = 97.3%) (Figure 15). Data from Somalia, derived from a single study, reported prevalence estimates of 17% using IgM and 21% using PCR.

**Figure 13.**
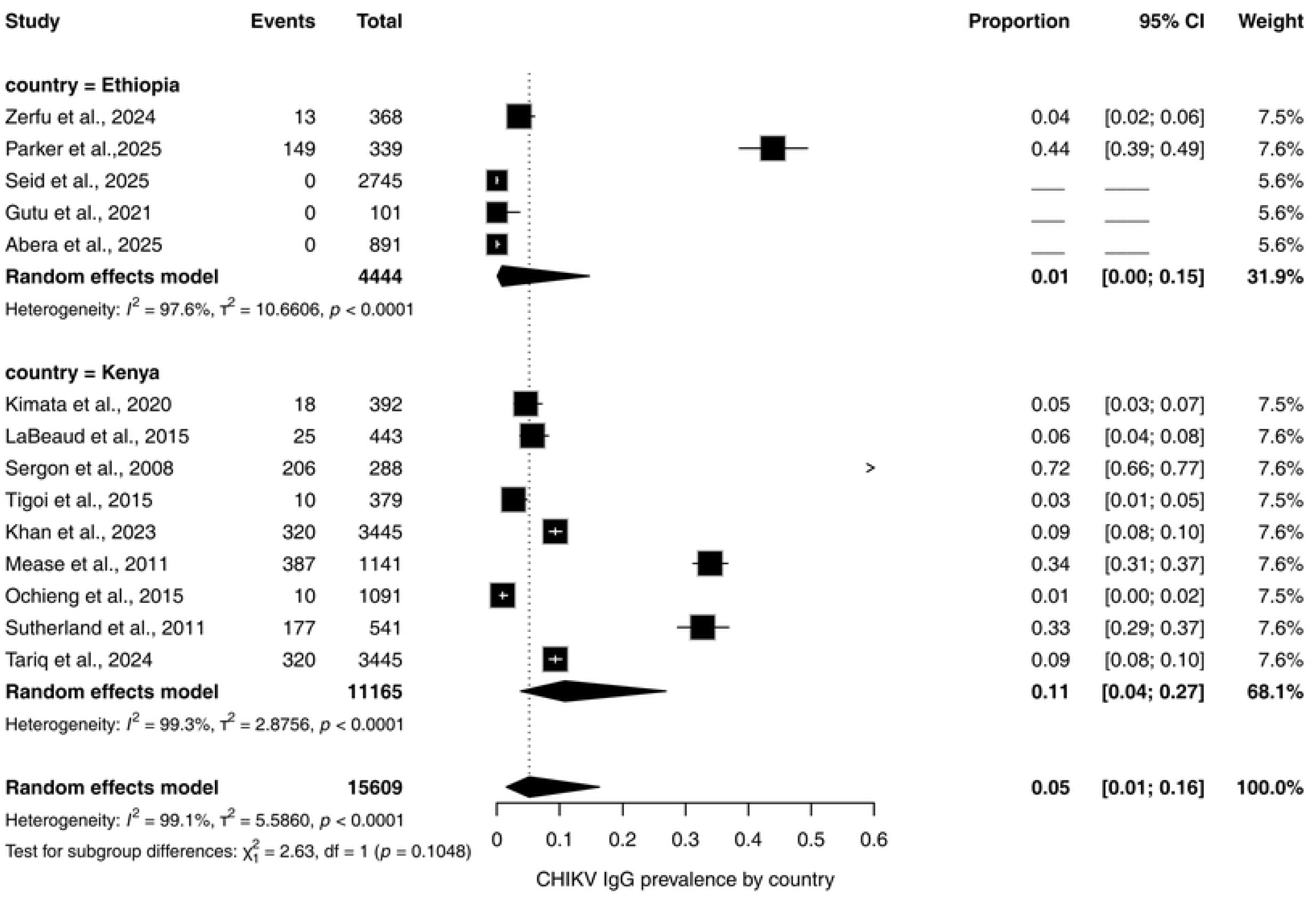
Forest plot showing the pooled prevalence of chikungunya virus IgG antibodies with subgroup analysis by country.

**Figure 14.**
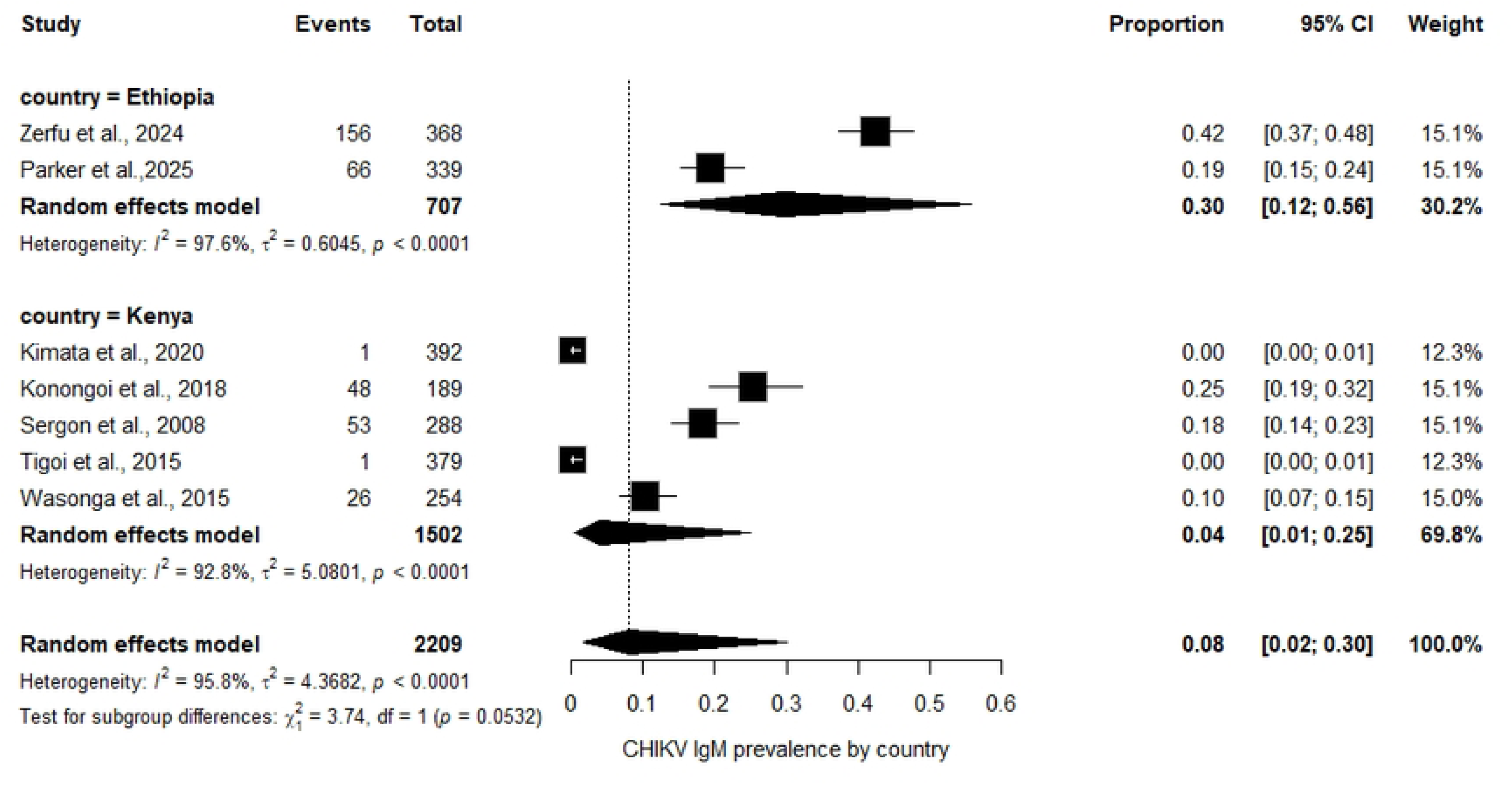
Forest plot showing the pooled prevalence of chikungunya virus IgM antibodies with subgroup analysis by country.

**Figure 15.**
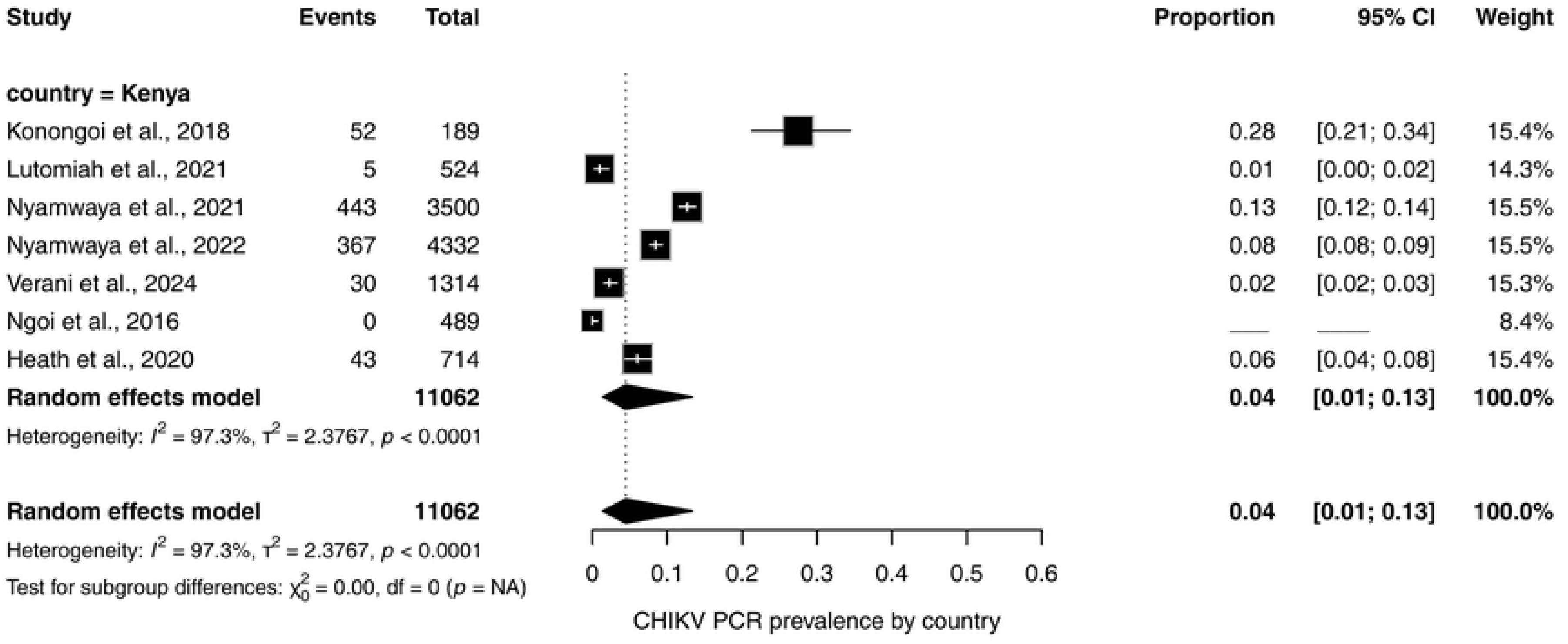
Forest plot showing the pooled prevalence of PCR-confirmed chikungunya virus infection with subgroup analysis by country.

### 3.8 Subgroup analysis of CHIKV prevalence

Subgroup analyses of CHIKV prevalence demonstrated variation across population groups and study settings. Prevalence was higher in mosquito vectors (6%) compared to febrile patients (3%); however, this difference was not statistically significant (χ² = 0.20, *P* = 0.6530) (Figure 16). In contrast, significant differences were observed across study settings (χ² = 17.06, *P* = 0.0002), with the highest prevalence reported in rural areas (15%), followed by peri-urban settings (3%), while interpretation of urban estimates was limited due to the availability of only one study. Across all subgroup analyses, substantial heterogeneity remained high (I² > 90%) (Figure 17).

**Figure 16.**
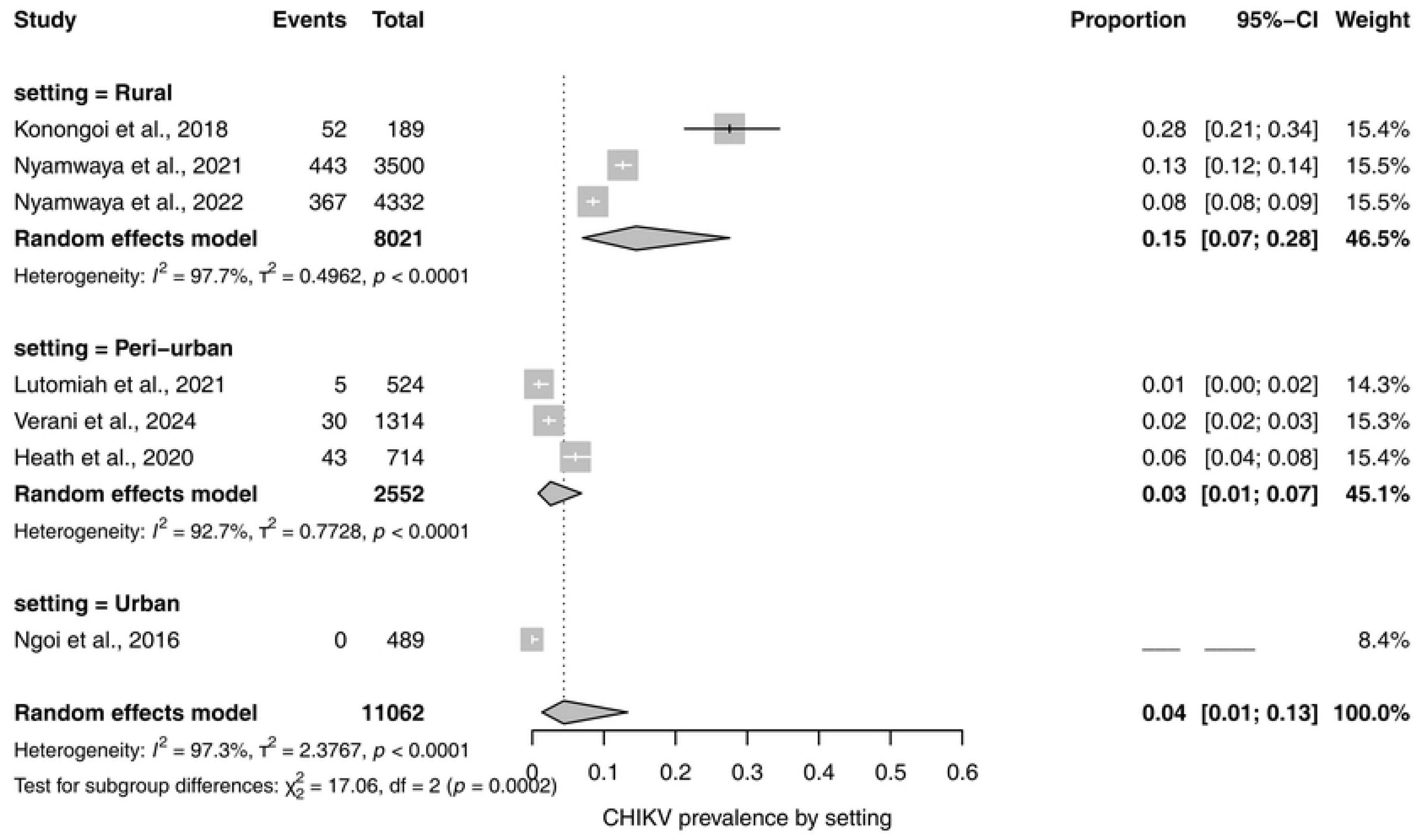
Subgroup analysis of PCR-confirmed chikungunya virus prevalence according to study population.

**Figure 17.**
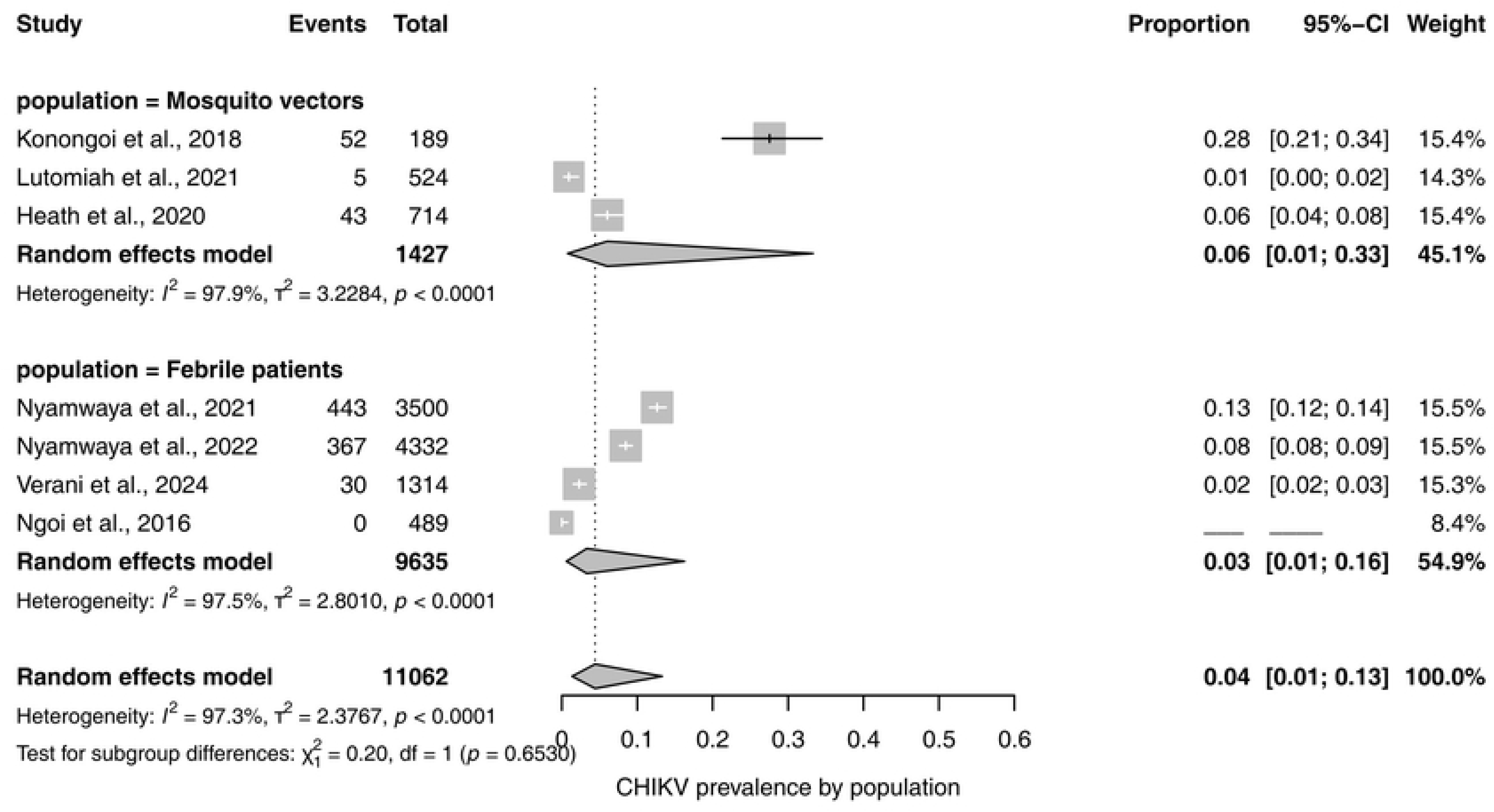
Subgroup analysis of PCR-confirmed chikungunya virus prevalence according to study setting.

### 3.9 Vector species reported in the included studies

Vector surveillance information was reported in 13 (20%) of the included studies. A diverse assemblage of mosquito genera associated with arbovirus transmission was identified across these studies (Figure 18). Mosquitoes belonging to the *Aedes* genus were the most frequently reported group overall, appearing in 92.3% of vector studies, followed by *Culex* species (53.8%) and *Anopheles* species (38.5%). Among the identified species, *Aedes aegypti* was by far the most reported vector. Several additional *Aedes* species were reported, including *Aedes africanus*, A*edes simpsoni, Aedes tricholabis, Aedes vittatus,* and *Aedes tarsalis.* Multiple *Culex* species, including C*ulex pipiens* and *Culex univittatus,* were also identified. Additionally, mosquitoes belonging to the genus *Anopheles*, including *Anopheles gambiae* and *Anopheles funestus,* were identified in several studies.

**Figure 18.**
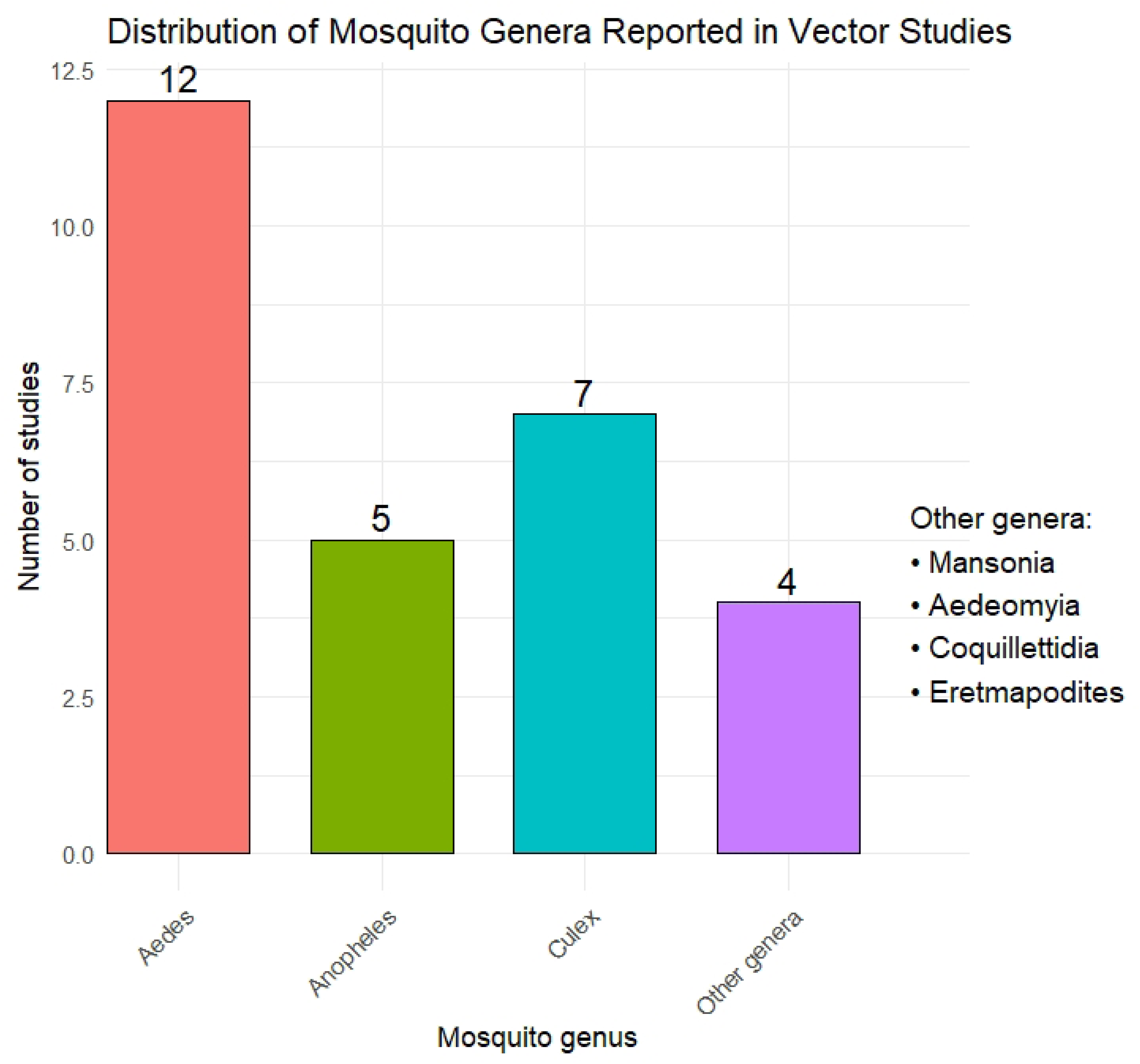
Frequency of mosquito genera and species reported in vector surveillance studies.

### 3.10 Research Landscape and Methodological Characteristics of Included Studies

The research landscape demonstrated that studies predominantly focused on disease prevalence and seroepidemiology (23/65, 35.4%), followed by outbreak investigation and epidemiology and clinical diagnosis, diagnostics, and surveillance, each accounting for 11 studies (16.9%). Risk factor assessment was the least represented research area, comprising only 3 studies (4.6%) (Figure 19).

**Figure 19.**
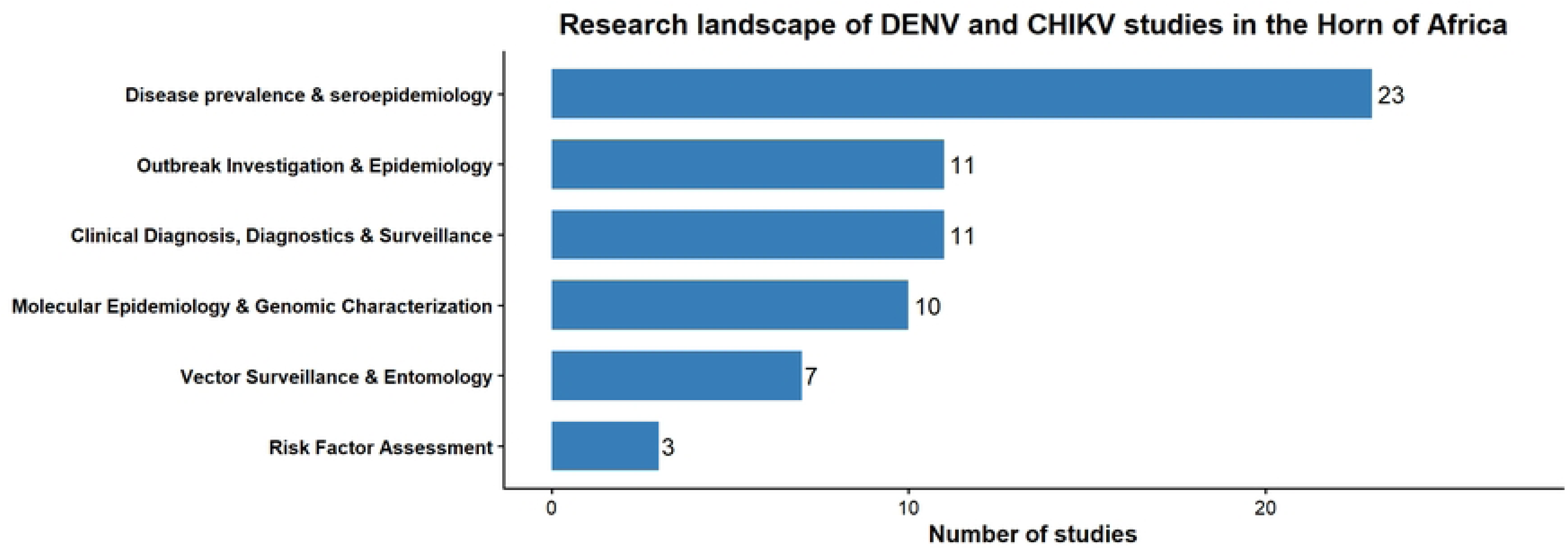
Distribution of research themes among the included studies.

Assessment of methodological limitations revealed that the most frequently reported challenges were sampling and representativeness (27/65, 41.5%) and diagnostic confirmation (27/65, 41.5%), followed by resource constraints (21/65, 32.3%) and serological limitations (19/65, 29.2%). Fewer studies reported limitations related to study design (13/65, 20.0%), insufficient epidemiological or vector data (11/65, 16.9%), and geographic and temporal coverage (10/65, 15.4%) (Figure 20).

**Figure 20.**
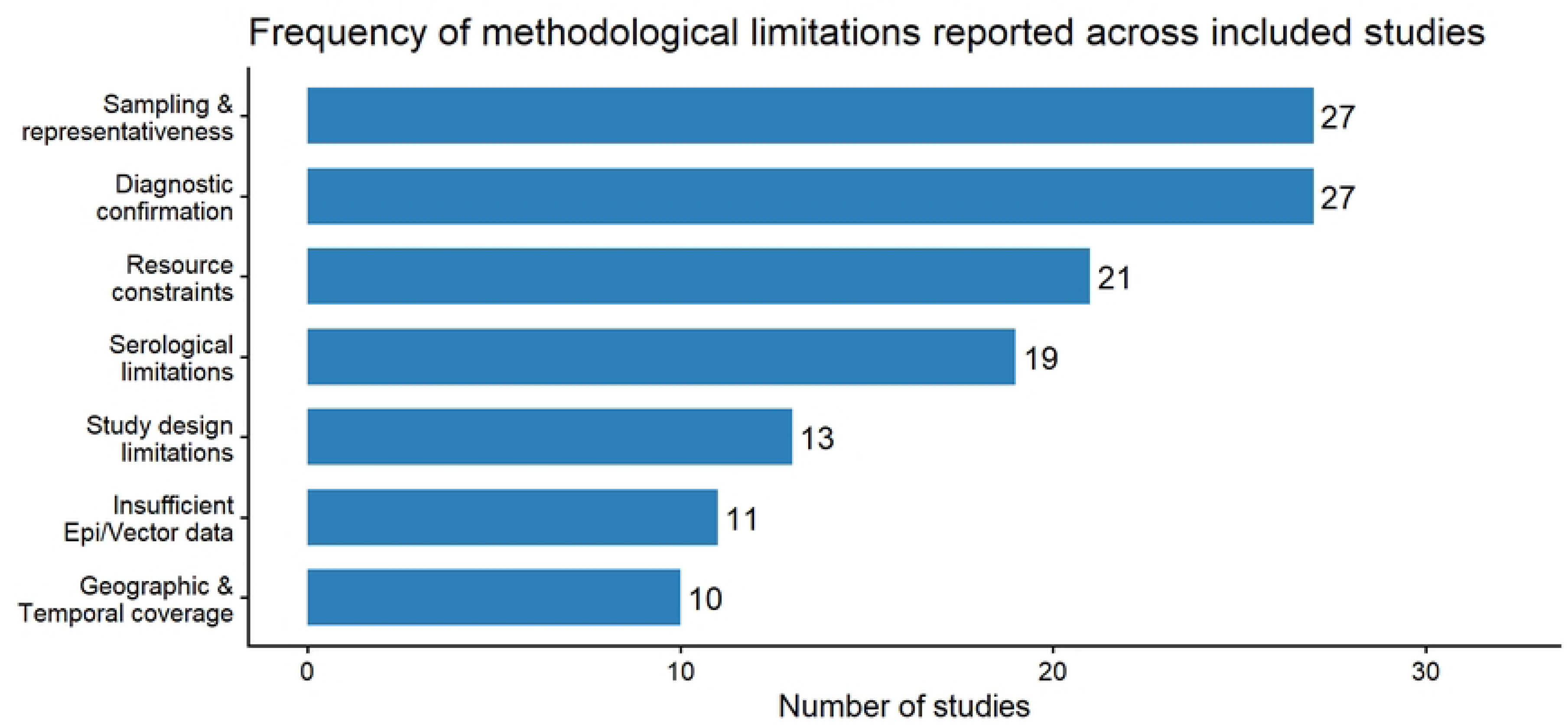
Reported methodological limitations among the included studies.

## 4. Discussion

This systematic review and meta-analysis synthesized findings from 65 studies to provide a comprehensive overview of DENV and CHIKV prevalence in the East African region, with a particular focus on Kenya, Ethiopia, and Somalia. The literature selection process identified a substantial number of records, with 65 studies ultimately meeting the inclusion criteria after rigorous screening and quality assessment.

This systematic review and meta-analysis shows that DENV and CHIKV represent a significant and growing burden in the Horn of Africa, supporting the recognition of the region as an emerging focus for arboviral transmission. The pooled DENV IgG seroprevalence for the region was 20% suggesting that an estimated 1 in 5 people in the region had previously been exposed to DENV, indicative of historic transmission. This estimate is similar to previous studies conducted in Sub-Saharan Africa with pooled prevalences of 25% and 24.8% [20,21]. Notably, however, the prevalence of the present study is much higher than the previous estimate of 3.6% that has been reported solely for Eastern Africa by Simo et al. (2019) [21] which indicates a substantial increase in DENV transmission in the Horn of Africa in recent years. Factors leading to this rise could be attributed to increasing urbanization, greater human mobility, climate and environmental changes, and the proliferation of *Aedes* mosquito populations across the region.

For CHIKV, significant differences were observed between countries and diagnostic markers. The estimated IgG seroprevalence of 11% in Kenya compares with other estimates in African settings including a pooled prevalence of 7.28% reported by Kayange et al. (2023) [22]. In contrast, the high prevalence (29%) of IgM in Ethiopia shows high burden of recent or active CHIKV transmission highlighting the increasing importance of Ethiopia as a potential hotspot for CHIKV circulation in the Horn of Africa. These findings are consistent with the pooled prevalence of 23.7% reported in a recent systematic review by Bangoura et.al (2024) [23]. Our findings collectively suggest that the transmission dynamics of CHIKV in the Horn of Africa are similar to those observed elsewhere in endemic tropical and subtropical settings with established competent *Aedes* vectors.

Our meta-analysis indicated widespread circulation of DENV and CHIKV in the Horn of Africa, and provided evidence for both historical widespread exposure and ongoing transmission. The pooled IgG seroprevalence of 20% for DENV indicates a significant level of previous exposure to the virus in the population. The lower pooled IgM prevalence of 10% indicates recent or current circulation of DENV. This pattern is consistent with the biological kinetics of dengue antibodies, where IgM becomes detectable approximately 5 days after fever onset and lasts for about 3 months, and IgG generally appears after 10 days and can persist for several years [24]. PCR-based CHIKV studies showed a pooled prevalence of 4% but a single study from Somalia reported significantly high prevalence estimates of 17% by IgM and 21% by PCR indicating active viral circulation during the study period. Collectively these results highlight the need for serological and molecular markers to be interpreted considering cumulative exposure, recent history of infection, and current dynamics of transmission in an effort to fully understand the epidemiology of dengue and chikungunya.

The variation in DENV and CHIKV prevalence across Kenya, Ethiopia and Somalia, and likely reflects epidemiological differences due to ecological, environmental and surveillance factors. The warm and humid conditions of the coastal lowlands favor the breeding of *Ae. aegypti*, while highland and arid areas generally show lower vector densities due to less favorable climatic conditions and limited breeding habitats [25]. Rapid urbanization, poor water supply, poor sanitation, and ineffective waste management, particularly in urban and peri-urban settings, further enhance vector proliferation and sustained transmission [26]. In addition, varying surveillance system and study designs influence the reported prevalence estimates, with community based serosurveys reflecting cumulative exposure and health facility based studies mostly reflecting recent and/or acute infections. Finally, regional differences in transmission dynamics are probably associated with variation in past outbreak history and population immunity, with higher seroprevalence in recent outbreak areas reflecting previous exposure [23].

We found considerable heterogeneity in DENV serotype distribution in the Horn of Africa. Overall, the most common serotype was DENV-2 16 (43.2%), followed by DENV-3 37.8%, DENV-1 6 (16.2%) and DENV-4 8.1%. This is important because DENV-2 has frequently been linked to severe clinical manifestations and has been involved in several large dengue epidemics globally [27]. The distribution of particular serotypes in different countries is likely due to a complicated interaction of factors including viral evolution, vector competence, host immunity and environmental factors that influence the transmission success of particular viral lineages [28]. DENV-2 and DENV-3 were the dominant serotypes in Kenya, while in Ethiopia DENV-3 was the most frequently reported serotype. DENV-2 and DENV-3 were only identified in Somalia. Significantly, the presence of all four DENV serotypes in Kenya and Ethiopia indicates increasing viral diversity and active local transmission in the region. Co-circulation of multiple serotypes is of particular concern for public health, as secondary infection with a different serotype may increase the risk of severe dengue through antibody-dependent enhancement [29]. Such multi-serotype circulation patterns have been reported in other dengue-endemic regions and are frequently associated with recurrent outbreaks and an increased burden of severe disease. In contrast, the low serotype diversity in Somalia is probably a result of the surveillance gaps, limited molecular characterization, and the small number of studies rather than the true absence of additional serotypes [15]. Together, these results highlight the need to improve molecular and genomic surveillance across the Horn of Africa to better monitor serotypes, prepare for outbreaks and develop evidence-based dengue prevention and control strategies [30].

The dominance of *Ae. aegypti* in the studies reviewed confirms it as the main vector of DENV and CHIKV transmission in the Horn of Africa. Its anthropophilic feeding behavior, adaptation to urban and peri-urban environments and preference for artificial water containers facilitate sustained transmission, especially in areas undergoing rapid urbanization, poor water management and sanitation. These findings are consistent with earlier studies identifying *Ae. aegypti* is the major vector of dengue and chikungunya worldwide [3,31]. *Culex* and *Anopheles* species were also reported, although there is no current evidence suggesting a significant role of these genera in DENV or CHIKV transmission in the region. However, detection of these species emphasizes the need for ongoing entomological surveillance and integrated vector management to monitor changing vector ecology and transmission dynamics.

The findings of this review have important implications for arboviral control in the Horn of Africa. The high burden of DENV and CHIKV disease, and evidence for active transmission and co-circulation of multiple DENV serotypes, suggests the need for these infections to be incorporated into routine surveillance systems and differential diagnosis of acute febrile illness. Key actions include improving access to affordable molecular and serological diagnostics, strengthening laboratory capacity, increasing clinical awareness and implementing integrated vector management, which includes community-based source reduction, improved water and waste management and targeted vector control. Genomic surveillance must be expanded to track viral evolution, identify new strains and improve preparedness for outbreaks. Current surveillance limitations, especially in Somalia and Ethiopia, demonstrate the need for regional collaboration and harmonised One Health surveillance.

The review also points to major research priorities. Most of the available studies were cross-sectional and conducted in Kenya, limiting understanding of disease incidence, seasonal patterns and transmission dynamics at sub-national levels. Aedes mosquitoes are the main vectors for the transmission of arboviruses, but there are few vector surveillance and ecological studies. Many studies suffered from non-representative sampling and lack of access to confirmatory laboratory diagnostics, which increased the risk of underdiagnosis and misclassification. Future studies should therefore not be confined to descriptive prevalence studies but should incorporate longitudinal and multidisciplinary approaches, combining epidemiological, clinical, entomological, environmental, molecular and genomic data, using standardized protocols to improve One Health surveillance and outbreak preparedness.

These results should be interpreted with several limitations in mind. The pooled prevalence estimates are limited in their comparability and generalizability due to high heterogeneity between studies, differences in diagnostic methods and predominance of cross-sectional designs. The geographic distribution was also uneven with relatively few studies from Somalia and parts of Ethiopia. Publication bias and overrepresentation of outbreak investigations may have overestimated prevalence estimates. Limited longitudinal and individual-level data also precluded detailed assessment of temporal trends, risk factors and clinical outcomes. Pooled estimates were based on individual studies rather than standardized national surveillance systems, and the true burden of DENV and CHIKV infections in the Horn of Africa is likely to remain underestimated.

## Conclusion

In conclusion, the systematic review and meta-analysis demonstrates that DENV and CHIKV are important and likely underrecognized arboviral threats in the Horn of Africa. Evidence of substantial seroprevalence, active viral circulation, co-circulation of multiple DENV serotypes, and widespread Aedes vectors indicates that these viruses are well established in the region. The findings emphasize the urgent need for strengthened surveillance systems, improved molecular and serological diagnostic capacity, sustained integrated vector control strategies, and enhanced regional collaboration to support early outbreak detection and effective public health response. Without coordinated intervention efforts, the burden of dengue and chikungunya in the Horn of Africa is likely to continue increasing, posing significant challenges for public health systems in the region.

## Data Availability

All data underlying the findings of this systematic review and meta-analysis are fully available without restriction. The extracted study-level dataset and risk of bias assessments are provided as Supporting Information files accompanying this article.

## Supplementary materials

Supplementary Table S1. Quality assessment of the studies included in the systematic review using the Joanna Briggs Institute (JBI) critical appraisal checklist.

## Authors’ contributions Declaration of Competing Interest

The authors declare that there is no conflict of interest.

## Ethics approval and consent to participate

Not applicable.

